# Changes in household purchasing of soft drinks following the UK Soft Drinks Industry Levy by household income and composition: controlled interrupted time series analysis, March 2014 to November 2019

**DOI:** 10.1101/2023.11.27.23299070

**Authors:** Nina T. Rogers, Steven Cummins, David Pell, Harry Rutter, Stephen J. Sharp, Richard Smith, Martin White, Jean Adams

## Abstract

**Objectives:** To examine changes in volume of and amount of sugar in purchases of soft drinks according to household income and composition, at 19 months following the implementation of the UK Soft drinks industry levy (SDIL).

**Design:** Controlled interrupted time series analysis

**Setting:** Representative households (mean weekly number of households =21,908) across Great Britain

**Participants:** Members of the Kantar Fast Moving Consumer Goods panel, a market research panel which collects data on weekly household purchases (eg: drinks, confectionery etc) between March 2014 to November 2019.

**Interventions:** The SDIL, is a two-tiered tax (announced in March 2016 and implemented in April 2018) on manufacturers of soft drinks. Drinks containing ≥8g sugar /100mls and ≥5 to <8g sugar/ 100mls are taxed at £0.24/litre and £0.18/litre, respectively. Soft drinks containing < 5g sugar/100ml are not subject to the levy. Levy exempt drinks, irrespective of sugar content, include milk and milk-based drinks, no-added-sugar fruit juice and powder used to make drinks.

**Main Outcome measures:** Absolute and relative differences in the volume of and amount of sugar in non-alcoholic soft drinks, confectionery and alcohol purchased weekly by household income (<£20,000, £20-50,000 or >£50,000) and composition (presence of children [<16years] in the household (yes or no), 19 months after SDIL-implementation, compared to the counterfactual scenario based on pre-announcement trends and using a control group (toiletries).

**Results:** By November 2019, overall purchased weekly sugar in soft drinks fell by 7.46g (95%CI: 12.05, 2.87) per household but volumes of drinks purchased remained unchanged, compared to the counterfactual based on pre-announcement trends. In low-income households, weekly sugar purchased in soft drinks decreased by 14.0% (95%CI: 12.1,15.9) compared to the counterfactual but in high income households increased by 3.4% (1.07,5.75). Similarly, among households with children, sugar purchased decreased by 13.7% (12.1, 15.3) compared to the counterfactual but increased in households without children by 5.0% (3.0,7.0). Low-income households and those with children also reduced their weekly volume of soft drinks purchased by 5.7% (3.7, 7.7) and 8.5% (6.8, 10.2) respectively. There was no evidence of substitution to confectionary or alcohol.

**Conclusion:** In the second year following implementation of the SDIL, there were sustained reductions in sugar derived from soft drink purchases, but no change in volume of soft drinks purchased. Effects on sugar purchased were greatest in those with the highest pre-SDIL purchasing levels (low-income households and those with children). The SDIL may contribute to reducing dietary inequalities.

**Trial registration:** ISRCTN18042742.

**Summary box:** *What is already known on this topic:* The World Health Organization recommends taxes on sugar sweetened beverages (SSBs) to improve population health; systematic reviews indicate these can successfully reduce population purchasing and consumption; differential impacts across demographic groups have been less studied. In the UK, SSB intake is highest in lower socioeconomic groups and children. The UK soft drinks industry levy (SDIL) successfully reduced household purchasing of sugary from soft drinks by a mean of 8.0g per household per week at one year; longer term and differential effects of across different demographic groups have not been studied.

*What this study adds:* 19 months following implementation of the SDIL, there were sustained reductions in sugar from purchased soft drinks of 7.5g per household per week, but no change in the volume of purchases suggesting the SDIL may lead to long-term health gains without harming industry. Households with the lowest incomes (<£20,000/year) had the largest reductions in purchases of sugar from soft drinks which (compared to pre-announcement trends), dropped by an average of 70g of sugar per household per week, equivalent to just over two 250ml servings of a drink containing 5g sugar per 100 ml per person per week; households with children living in them reduced their purchasing of sugar from soft drinks by 56g per household per week. The SDIL may contribute to reducing existing inequalities in dietary intake.

## Introduction

Consumption of sugar sweetened beverages (SSBs) is associated with poor health outcomes including non-communicable diseases such as cardiovascular disease[1], type II diabetes[1], obesity[2–5] and dental caries[6]. There are inequalities in consumption of SSBs with lower socio-economic groups consuming more [7,8]. High intake of SSBs is also common among children and adolescents [9] and is linked to overweight and obesity in this age group[10].

The introduction of SSB taxes in a number of countries has been seen as largely successful as a measure to support reductions in dietary intake of added sugar via SSBs[11–13]. Indeed, the World Health Organization has recommended taxation of SSBs to reduce consumption of added sugars and improve health[14,15]. In response to the UK childhood obesity crisis, the UK Soft Drinks Industry Levy (SDIL) on manufacturers, importers and bottlers of soft drinks was announced in March 2016 and implemented in April 2018[16]. This differed from most other SSB taxes as its primary aim was to incentivise reformulation, rather than to pass higher prices of soft drinks to consumers[17]. The SDIL was designed as a two-tiered levy with a higher tier for drinks containing over 8 g of sugar per 100ml (levied at a rate of £0.24 per litre) and a lower tier for drinks containing 5-8 g of sugar per 100 ml (levied at a rate of £0.18 per litre)[16]. Drinks with less than 5g sugar per 100ml are not levied[16]. A number of categories are exempted and not levied irrespective of sugar content, e.g. no-added-sugar fruit juices, milk based drinks, and drinks sold as powder. Companies manufacturing less than one million litres/year are also exempt. One year after the implementation of the UK SDIL, households in the UK were purchasing 2.7% less sugar from take-home drinks (compared to the pre-announcement period) while the volumes purchased had increased by 2.6%[18], suggesting reformulation of SSBs had occurred – a finding reinforced by analyses of the sugar content of drinks available in UK supermarkets [19].

However, while evidence suggests that SSB taxes have been effective at reducing sales and dietary intake of added-sugar from SSBs, it is uncertain whether they reduce inequalities in sugar consumption from SSBs[11]. In the UK, no study has examined the effect of the SDIL across sociodemographic groups. This is an important gap because in high-income countries, such as the UK, the burden of obesity and other diet-related NCDs disproportionately affects those with lower educational attainment [20], lower income[21] and those living in deprived neighbourhoods[22]. Children have been identified as a particularly important target population for obesity prevention measures. While microsimulation modelling studies have projected similar health benefits across socioeconomic groups [23,24] or greater health benefits for health in lower income groups [25–27] only a few real-world studies have studied these effects. These report mixed findings. In Chile, Catalonia and Philadelphia higher socioeconomic groups were more responsive to SSB taxes [12,28,29]. However, in Mexico, Tonga and elsewhere in the USA, lower socioeconomic groups were more responsive [30–33]. These differences in response to SSB taxes across socioeconomic groups might reflect the structure of differing SSB taxes, different background contexts as well as differences in particular outcomes studied – including sales, purchasing and expenditure. Fewer studies have explored differences in the effect of SSB taxes in children vs adults, but one study from Mexico found greater impacts in households with children than without [31].

Furthermore, evidence on the long-term (>12 months) impacts of SSB taxes on consumption of soft drinks is scarce. However, sustained reductions in purchases of SSBs have been observed in Mexico two years after the tax was implemented and compared to pre-tax trends[34] with a suggestion of some plateauing in purchasing by the third year [35].

To add to this evolving literature, we use controlled interrupted time series (CITS) analysis to extend earlier analyses of overall effects to nineteen months post implementation; and determine whether UK household purchases of sugar in, and volume of, soft drinks changed according to household income levels and in households with and without children, following the announcement and implementation of SDIL. We also examine if there is any evidence of substitution occurring by examining changes in purchases of sugar from confectionery or volume of alcohol.

## Methods

### Study timeline

CITS analysis was used to compare changes in the amount of sugar in, and volume of, purchased soft drinks bought for consumption in the home, examining the effects of both the announcement and the implementation of the SDIL, with the counterfactual scenario in which neither the announcement nor implementation happened. The CITS ran from week one in March 2014, through the time of the SDIL announcement (March 2016; study week 108), and the SDIL implementation [36] (April 2018; study week 214) until its final week in November 2019 (study week 295).

### Data Source

We used data from Kantar Fast Moving Consumer Goods (KFMCG) panel, a market research company which collects household panel data on purchases of food, drink and other items from households in Great Britain (thus excluding Northern Ireland). KFMCG provided household purchasing data at the weekly level. The weekly mean number of households was 21,908. Households recruited into the panel are given a handheld scanner to record the barcodes of purchased items brought into the home and a book of barcodes to record unpackaged items. The information (including online sales and deliveries) is uploaded and sent to KFMCG who link the purchasing information to nutritional data on a continual basis. Households record and update their demographic characteristics annually and as an incentive for taking part they receive gift vouchers equivalent to £100 ($122; €112) annually. KFMCG excludes households that record fewer than six purchases weekly along with those whose adjusted weekly spend is lower than an undisclosed minimum.

### Product categories

Purchased soft drinks considered in the study included both levy-liable and levy-exempt types that were purchased and brought into the home. Inclusion of both levy-exempt and levy-liable soft drinks in the study enabled examination of the full impact of the SDIL on all soft drink purchases and captures potential soft drink products that may have been used as substitutes but not otherwise included if levy-exempt soft drinks were not considered in the analysis. In sensitivity analyses, purchases of alcohol (including alcoholic and alcohol replacement drinks) and confectionery (sugar and chocolate confectionery) were explored separately to determine whether any reductions in sugar from, or volumes of, soft drink purchases were substituted by increases in purchasing of alcohol or sugar from confectionery. To account for background trends in household purchases, toiletries (shampoo, hair conditioner, and liquid soap) was incorporated as a non-equivalent control category.

### Household demographics

Total gross household income was categorised into three groups, less than £20,000 (low), £20,000-49,999 (middle) and £50,000 and over (high). Median annual household income in the UK in 2019 was estimated to be ∼ £45,000[37]. Households were categorised into those with children aged less than 16 years present and those without.

### Statistical analysis

Prior to analysis, products were assigned to the SDIL relevant groups (e.g. all soft drinks, alcohol, confectionery and toiletries) based on product groups assigned by KFMCG and product names. Analysis was based on weekly lists of purchasing by product line, which report the type of purchase, sugar content (per 100 g/ml) and volume or mass purchased. Proprietary grossing up weights, created by Kantar Worldpanel, were used throughout our analysis to extrapolate from the size of the panel to the size of the population in Great Britain (GB) and to ensure the sociodemographic spread of the panel was representative of the GB population. Weekly household sugar purchases were calculated as sum of all (sugar concentration * volume * KWP weight)/number of households. In subgroup analysis, weekly purchasing within a demographic group was further adjusted by multiplying it by the proportions of households from the population of Great Britain that were in each demographic group [38,39]

CITS was performed using a controlled generalised least squares model with an autocorrelation-moving average (ARMA) correlation structure where the autoregressive order (p) and moving average order (q) were selected to minimise the Akaike Information Criterion (AIC) value of the model. All models included adjustment for mean monthly temperature and the months of December and January, since purchasing of soft drinks is influenced by seasonal factors (see supplementary material). Predicted counterfactual values (assuming the SDIL had neither been announced nor implemented) were calculated from the model. The difference in weight or volume between the observed and counterfactual values was estimated at week 295 (03/11/2019) and expressed in absolute grams or mls and as a percentage. Confidence intervals in this study were calculated from standard errors estimated using the delta method [40] Analysis was conducted in R version 4.1.0.

#### Changes to Protocol

Three changes were made to the published protocol[41]. Firstly, KFMCG provided weekly rather than monthly purchasing data which allowed us to improve the precision of our findings. Secondly, we initially proposed the CITS to finish in March 2020, two years after SDIL was implemented. However, because of potential household stockpiling of grocery products in anticipation of (i)the UK leaving the European Union in December 2019 and (ii) national lockdown due to the Covid-19 pandemic [36] follow-up was ended in November 2019. Thirdly, to examine disparities across socioeconomic groups, socio-economic position was operationalised as household income, which was considered a stronger indicator of material living standards, compared to social class of the main household member.

#### Patient and public involvement

A steering group, including two lay members, meet twice a year to discuss the broader issues around SDIL evaluation. The public and participants were not involved in developing the research question or other aspects of the design reported here.

## Results

Table 1 summarises the mean volume and weight of sugar in drinks purchased per household in the week prior to the SDIL announcement and the week prior to its implementation, and in the final week of follow-up (19 months post implementation) in the total population, by income group and households with and without children. In all socio-demographic groups, average weekly sugar purchased in drinks reduced over the study period. In the week prior to the announcement, households in the lowest income group purchased nearly twice as much sugar from, and volume of, soft drinks than mid-income households and approximately four-times more sugar from, and volume of, soft drinks than households in the highest income groups. Households with children purchased approximately 40% more sugar and 30% higher volume of soft drinks than households without children.

**Table 1:**
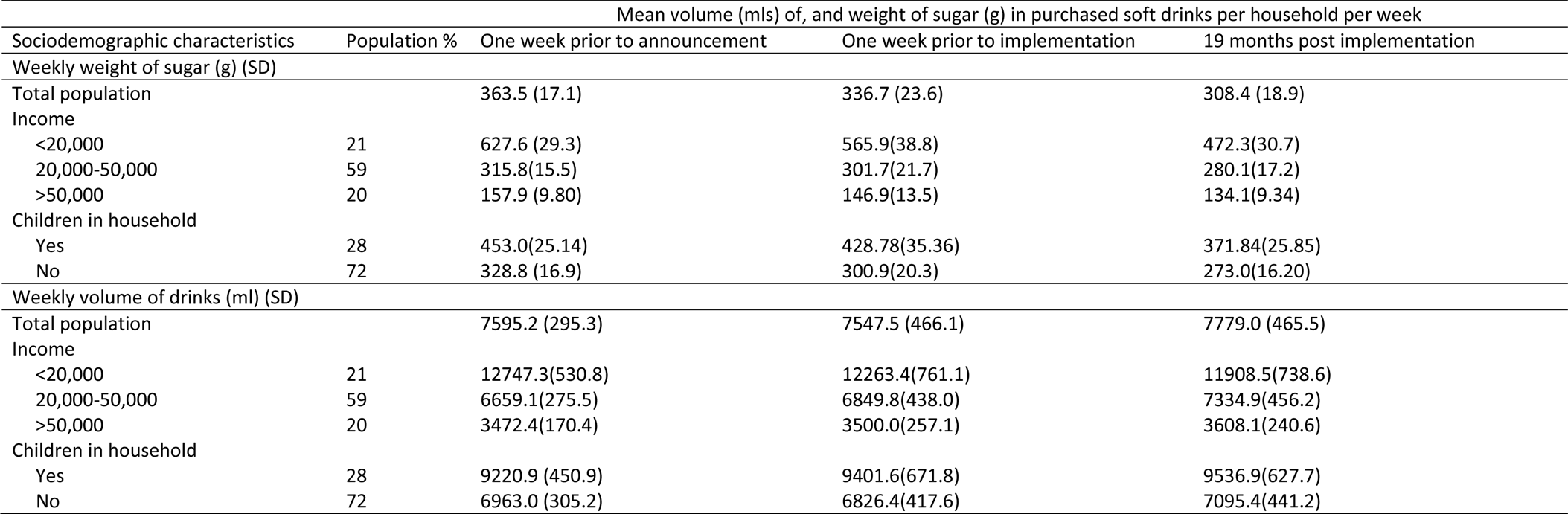
Mean weight of sugar in, and volume of purchased soft drinks per household per week in the week prior to announcement, implementation and 19 months post-implementation of the UK soft drinks industry levy.

Unless stated otherwise, all estimates below are per household per week, with respect to the counterfactual scenario (estimated from modelled pre-announcement trends (weeks 1-108) at 19 months post-implementation (November 2019 or time point week 295))

### Changes in amount of sugar from purchased soft drinks

Across all households in GB there was a 7.46g [95%CI: 2.87, 12.05] or 2.56% [95%CI:0.62, 4.49]) reduction in weight of sugar purchased from soft drinks (figure 1, table 2). The largest reduction was observed in the lowest income households (Figure 2) and households with children (Figure 3). Small increases in sugar purchased from soft drinks were seen in high income households and households without children. The sugar purchased from soft drinks was 70.27g [60.63, 79.91] or 13.98% [12.07,15.9] lower in low income households per household per week and 56.39g [49.82, 62.97] or 13.67% [12.08,15.27] lower in households with children at 19 months post-implementation compared to the counterfactual. Purchased sugar from soft drinks was 4.38g (1.37, 7.39) or 3.41% (1.07, 5.75) higher in high income households and 12.2g (7.29, 17.18) or 5.01% (2.99, 7.04) higher in households with no children present, respectively. Sugar purchased via soft drinks in middle-income households remained unchanged.

**Figure 1:**
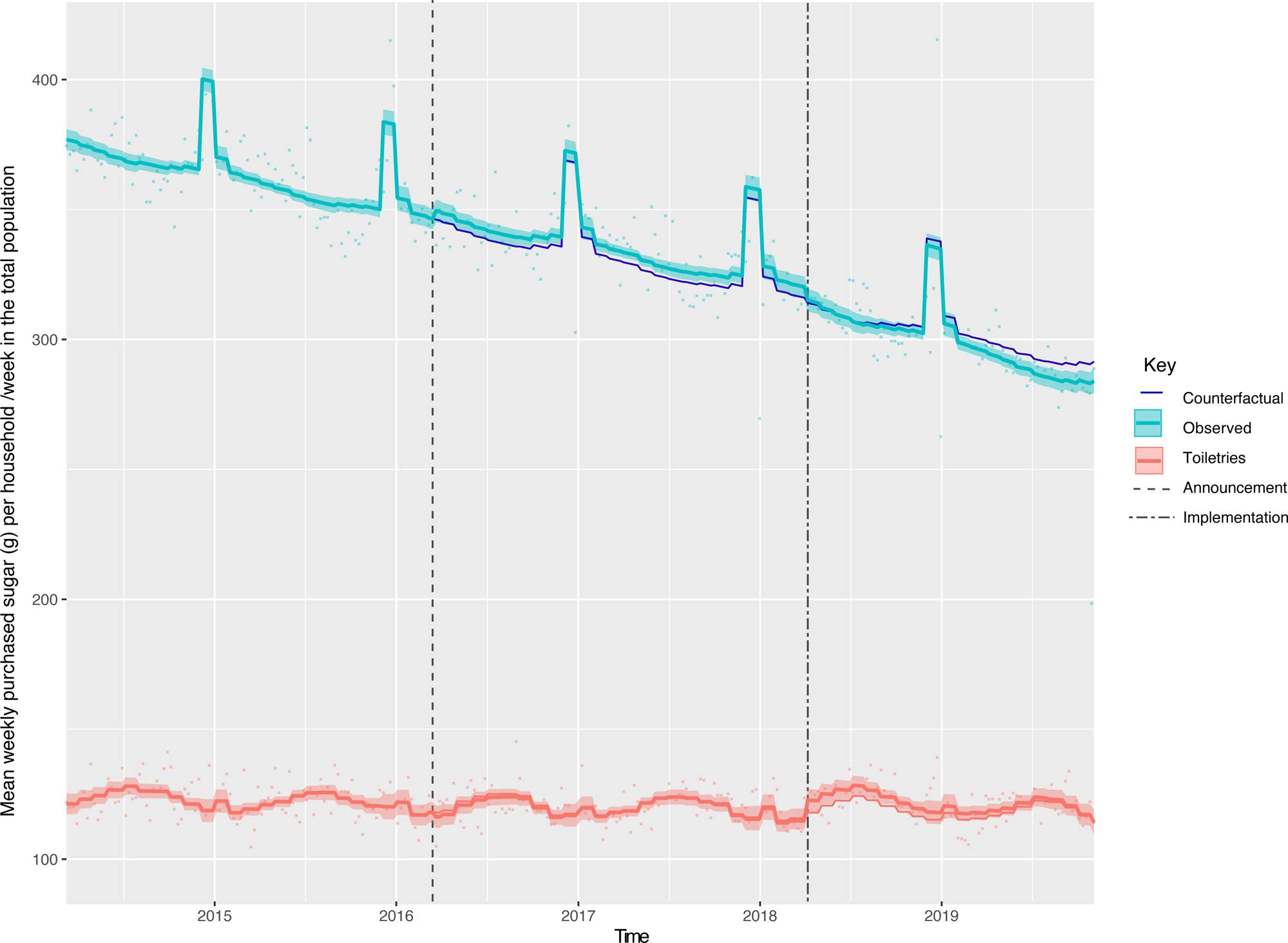
Weight (g) of sugar from soft drink products purchased per household per week in the total population, from March 2014 to November 2019. Observed and modelled amounts of sugar in all soft drinks (drinks liable to the SDIL and non-liable drinks). Light blue points show observed data and light blue lines (with light blue shadows) shows modelled data (and 95% confidence intervals) of sugar from purchased soft drinks. The dark blue line indicates the counterfactual line based on preannouncement trends and had the announcement and implementation not happened. The red line (and shadow) indicates modelled toiletries (control group). The first and second dashed lines indicate the announcement and implementation of SDIL, respectively.

**Figure 2:**
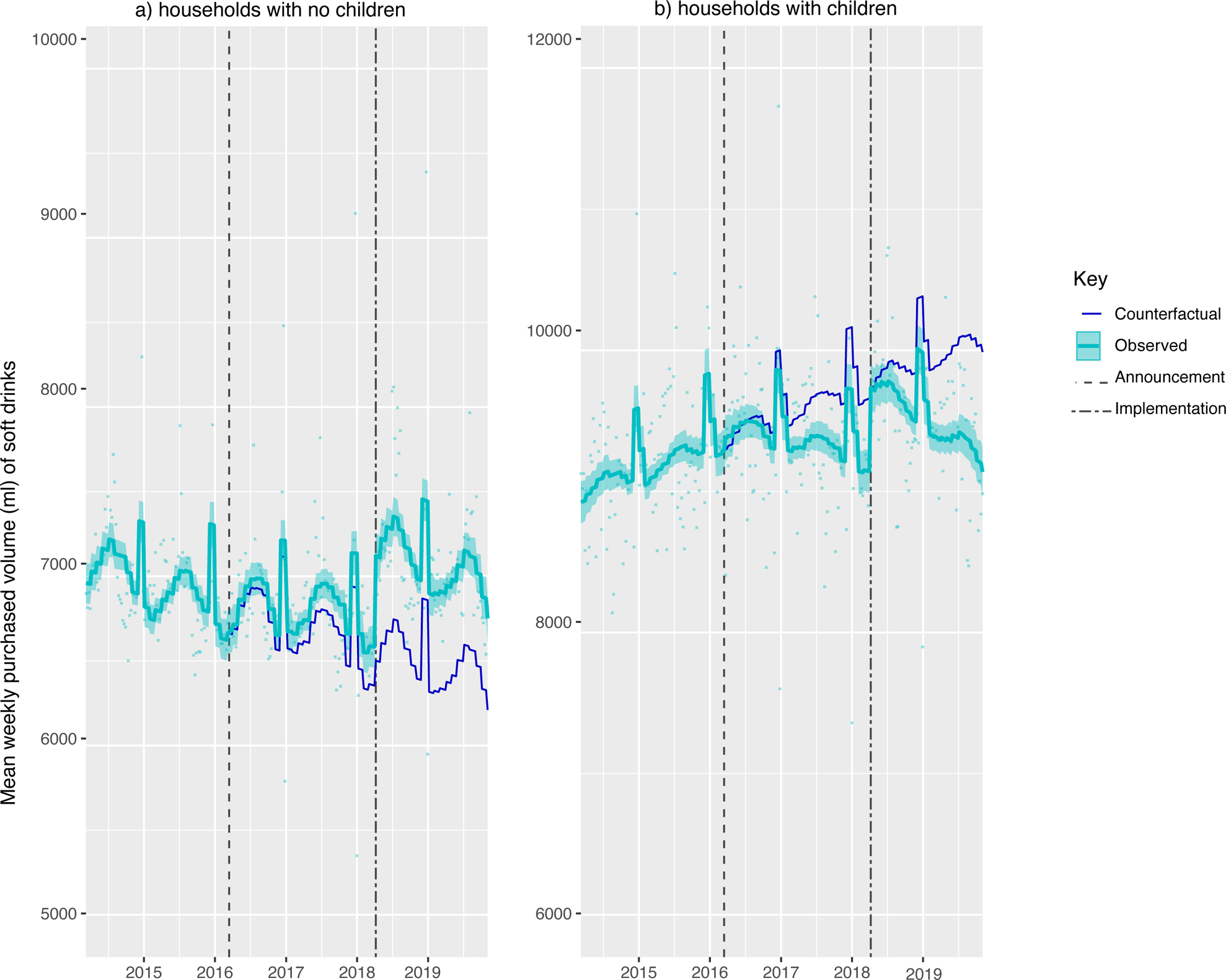
Weight (g) of sugar from soft drink products purchased per household per week, by gross household income levels, from March 2014 to November 2019. Observed and modelled amounts of sugar in all soft drinks (drinks liable to the SDIL and non-liable drinks) by annual gross household income levels of a) <20,000 b) £20,000-£50,000 and c) £50,000 or more. Light blue points show observed data and light blue lines (with light blue shadows) shows modelled data (and 95% confidence intervals) of sugar from purchased soft drinks. The dark blue line indicates the counterfactual line based on preannouncement trends and had the announcement and implementation not happened. The first and second dashed lines indicate the announcement and implementation of SDIL, respectively. The scales on the Y axis varies between panels and modelled toiletries have been removed (see figure S1 for inclusion of toiletries) to maximise the resolution of the graphs

**Figure 3:**
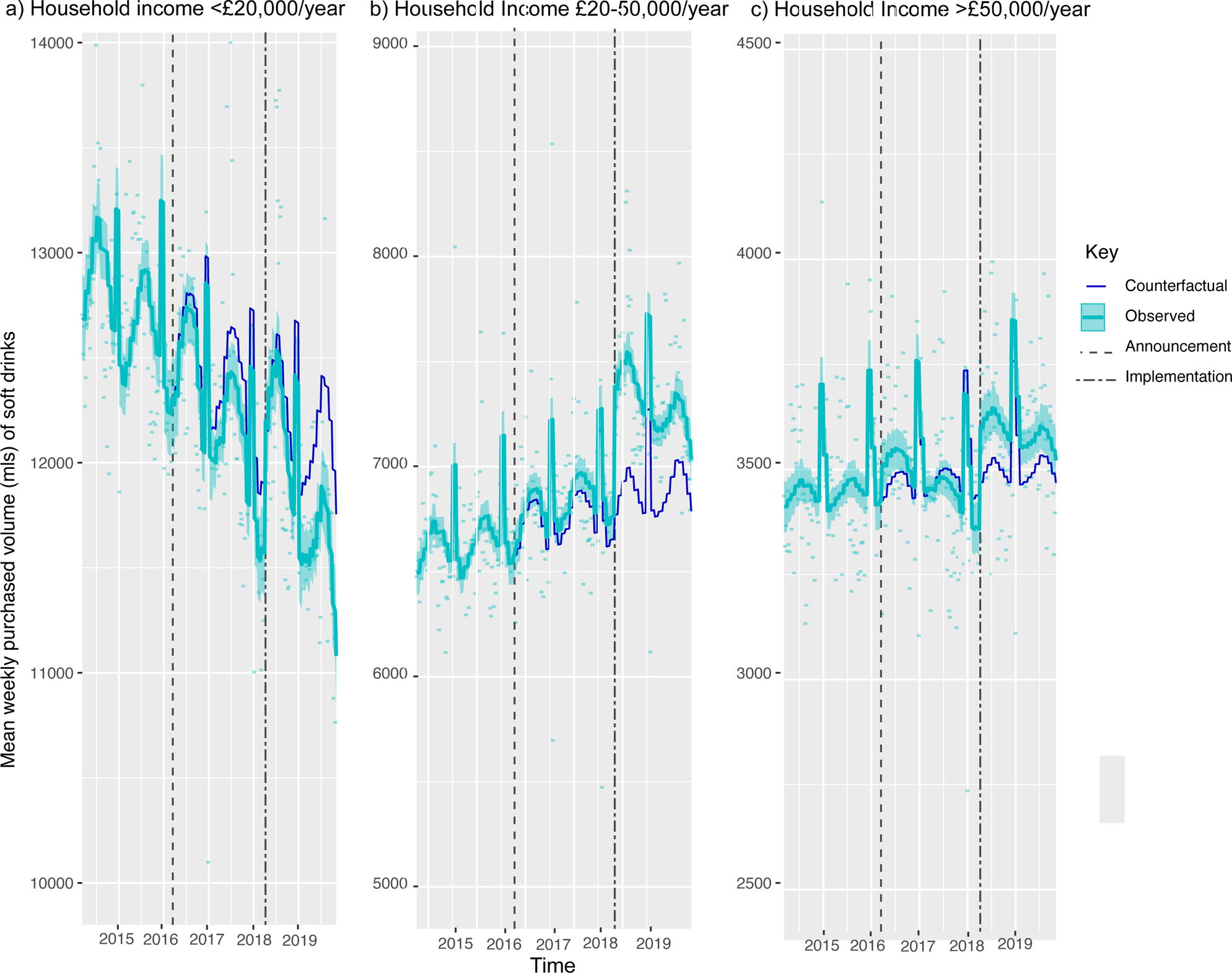
Weight (g) of sugar from soft drink products purchased per household per week, by whether households have children or not, from March 2014 to November 2019. Observed and modelled amounts of sugar in all soft drinks (drinks liable to the SDIL and non-liable drinks) by a) households with no children b) households with children (<16 years). Light blue points show observed data and light blue lines (with light blue shadows) shows modelled data (and 95% confidence intervals) of sugar from purchased soft drinks. The dark blue line indicates the counterfactual line based on preannouncement trends and had the announcement and implementation not happened. The first and second dashed lines indicate the announcement and implementation of SDIL, respectively. The scales on the Y axis varies between panels and modelled toiletries have been removed (see figure S2 for inclusion of toiletries) to maximise the resolution of the graphs

**Table 2:**
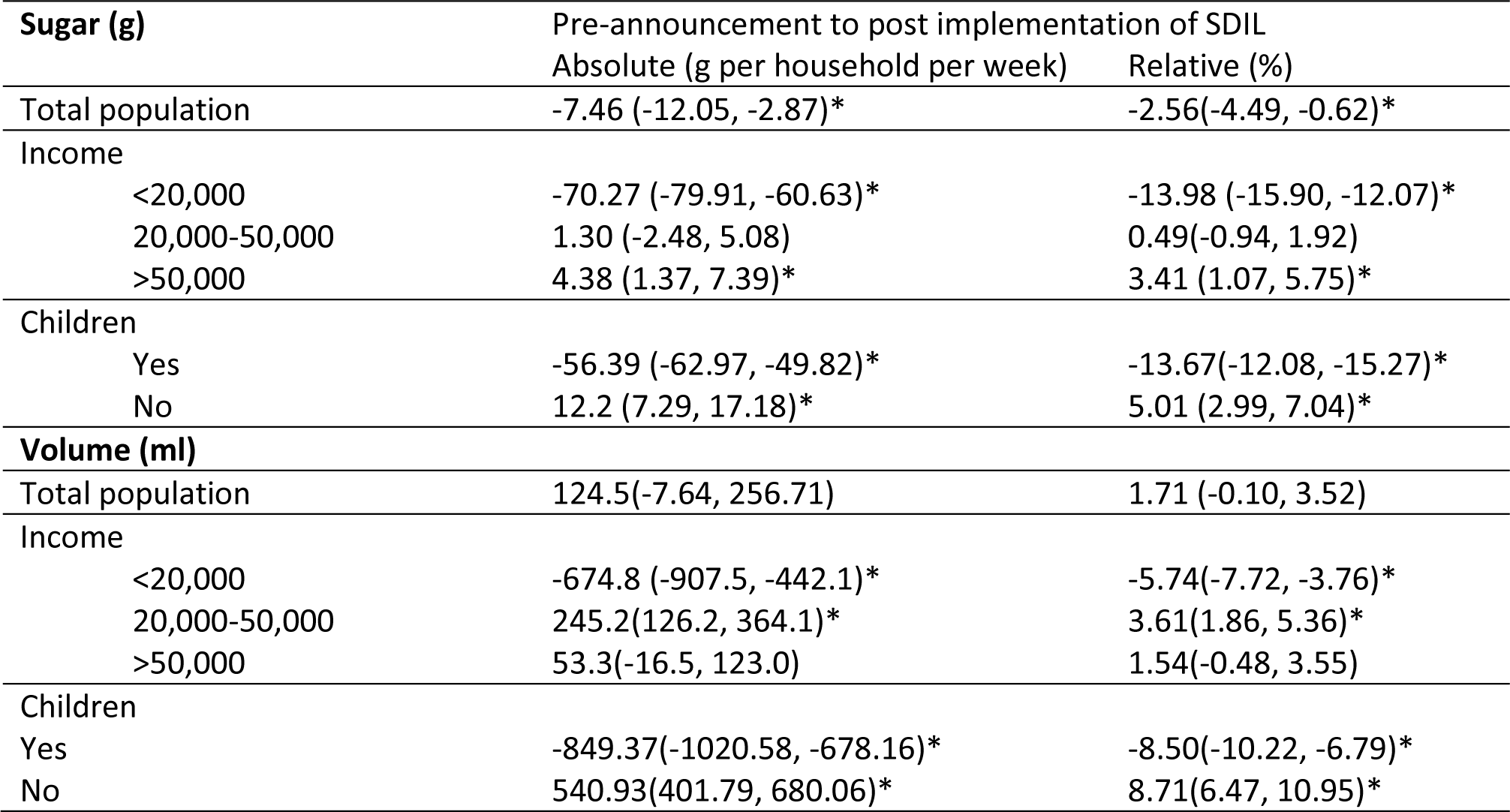
Absolute and relative changes in volume of, and weight of sugar in <u>soft drinks</u> purchased per household per week, compared to the counterfactual estimated from pre-announcement trends, at 19 months post-implementation of the UK soft drinks industry Levy.

### Change in volume of purchased soft drinks

Compared to the counterfactual, at 19 months post-implementation there was no overall change in the volume of soft drinks purchased across all households (figure 4, Table 2). However, there were reductions in the volumes of drinks purchased by the lowest income households (Figure 5) and those with children (Figure 6), with increases in middle-income households and households without children. The volume of drinks purchased was 674.8ml [442.1, 907.5] or 5.74% [3.76,7.72] lower in low-income households and 849.37ml (678.16, 1020.58) or 8.50% (10.22, 6.79) lower in households with children. In middle-income households and households without children the volume of drinks purchased was higher by 245.2ml (126.2, 364.1) or 3.61% (1.86, 5.36) and 540.93ml (680.06, 401.79) or 8.71% (6.47, 10.95), respectively.

**Figure 4:**
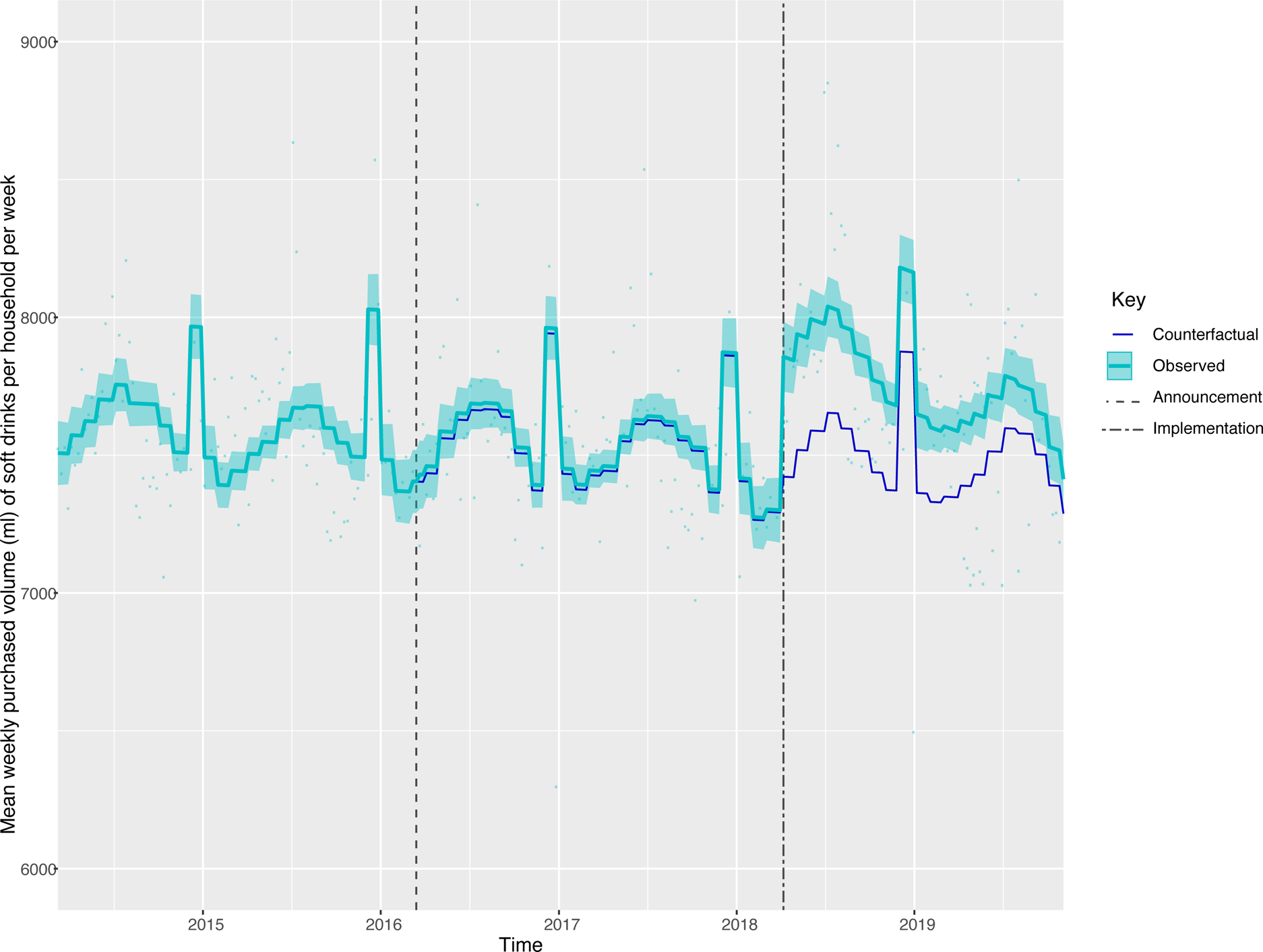
Volume (mls) of soft drink products purchased per household per week in the total population, from March 2014 to November 2019. Observed and modelled volumes of all soft drinks (drinks liable to the SDIL and non-liable drinks). Light blue points show observed data and light blue lines (with light blue shadows) shows modelled data (and 95% confidence intervals) of volumes of purchased soft drinks. The dark blue line indicates the counterfactual line based on preannouncement trends and had the announcement and implementation not happened. The first and second dashed lines indicate the announcement and implementation of SDIL, respectively. Modelled toiletries have been removed to maximise the resolution of the graphs.

**Figure 5:**
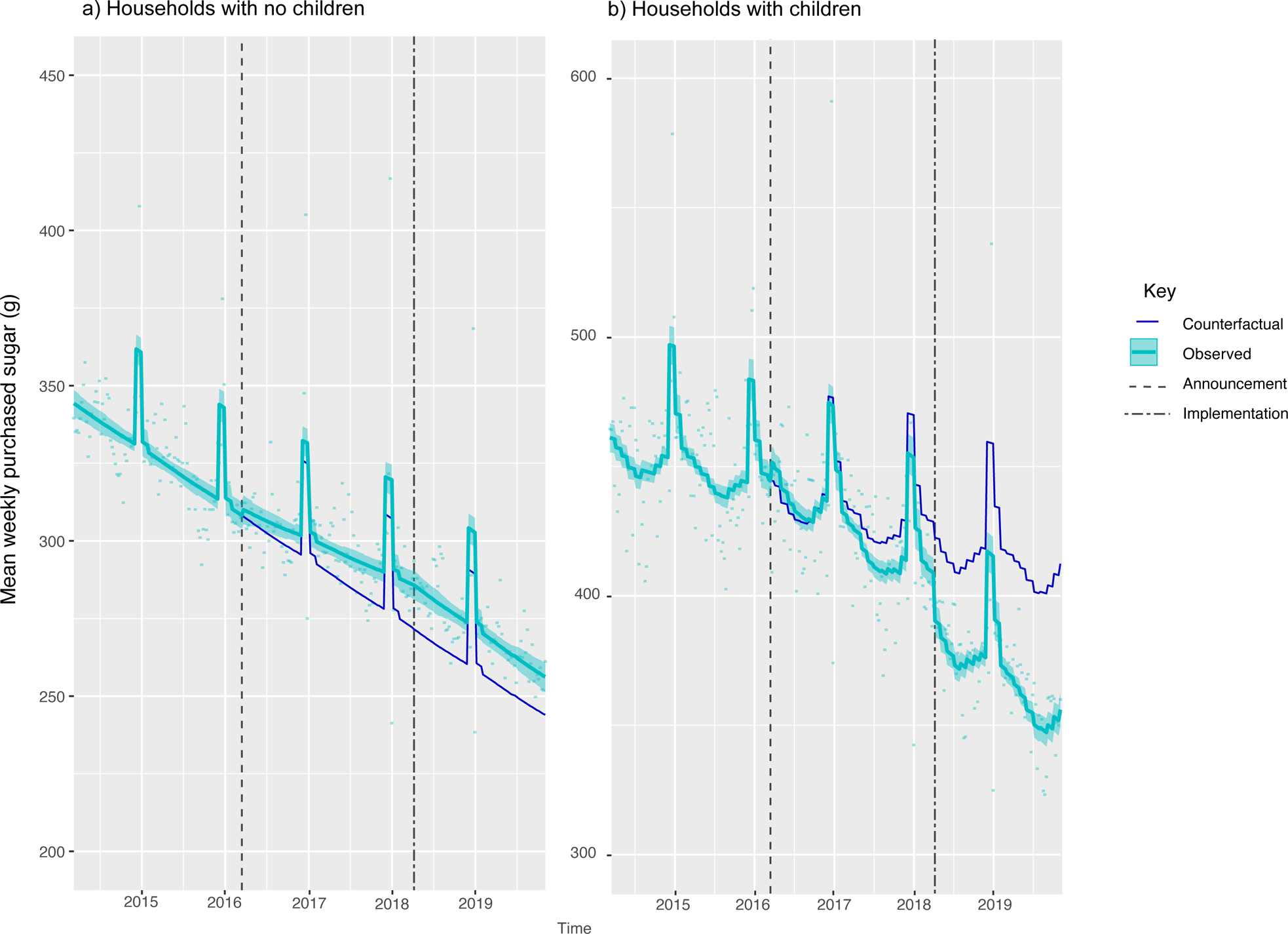
Volume (mls) of soft drink products purchased per household per week, by gross household income levels, from March 2014 to November. Observed and modelled volumes of soft drinks (drinks liable to the SDIL and non-liable drinks) by annual gross household income levels of a) <20,000 b) £20,000-£50,000 and c) £50,000 or more. Light blue points show observed data and light blue lines (with light blue shadows) shows modelled data (and 95% confidence intervals) of sugar from purchased soft drinks. The dark blue line indicates the counterfactual line based on preannouncement trends and had the announcement and implementation not happened. The first and second dashed lines indicate the announcement and implementation of SDIL, respectively. The scales on the Y axis varies between panels and modelled toiletries have been removed (see figure S3 for inclusion of toiletries) to maximise the resolution of the graphs

**Figure 6:**
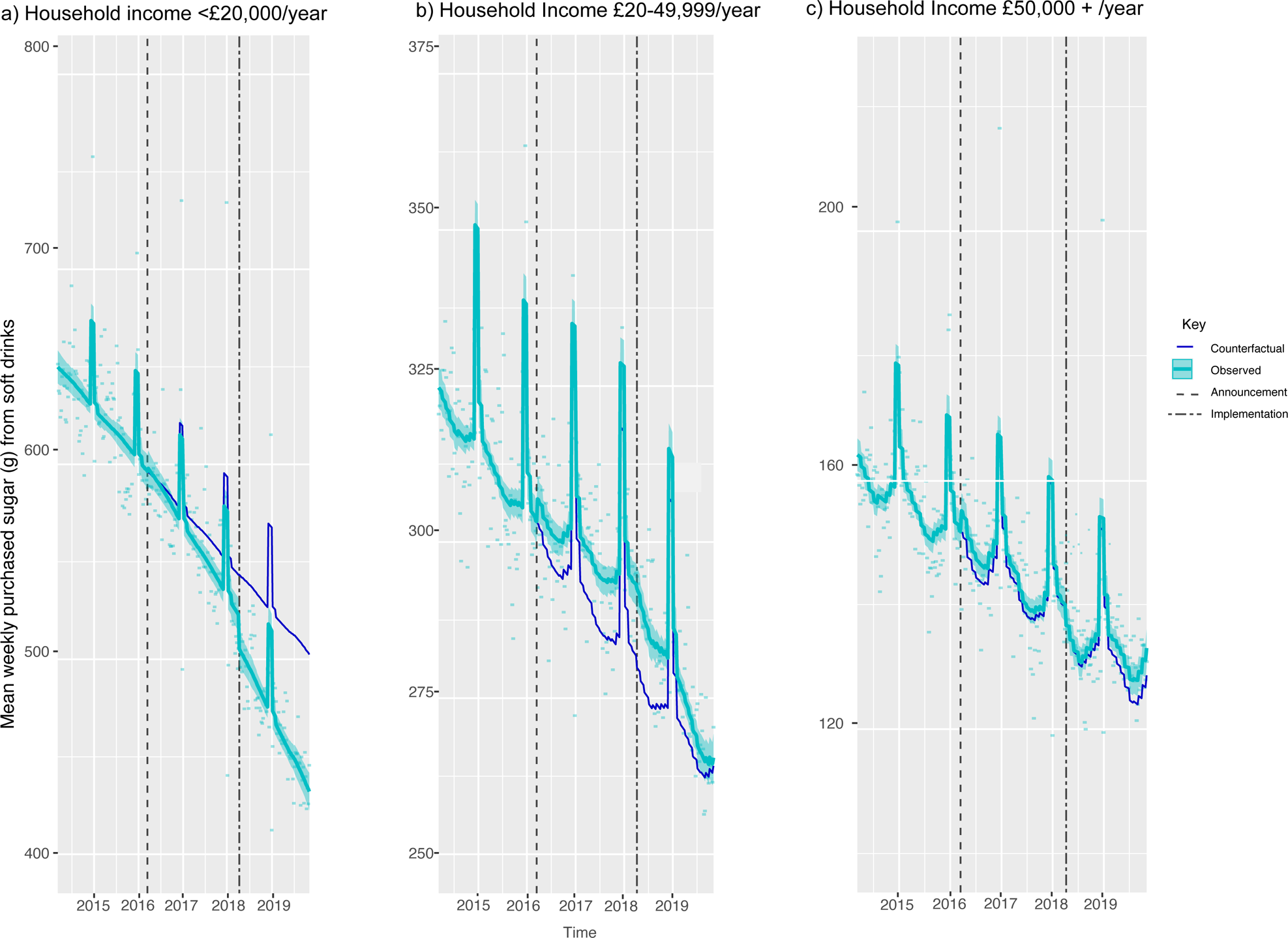
Volume (mls) of soft drink products purchased per household per week, by whether households have children or not, from March 2014 to November. Observed and modelled volumes of soft drinks (drinks liable to the SDIL and non-liable drinks) by a) households with no children b) households with children (<16 years). Light blue points show observed data and light blue lines (with light blue shadows) shows modelled data (and 95% confidence intervals) of sugar from purchased soft drinks. The dark blue line indicates the counterfactual line based on preannouncement trends and had the announcement and implementation not happened. The first and second dashed lines indicate the announcement and implementation of SDIL, respectively. The scales on the Y axis varies between panels and modelled toiletries have been removed (see figure S4 for inclusion of toiletries) to maximise the resolution of the graphs

### Purchasing of sugar through confectionery

Purchasing of sugar via confectionery was unchanged across all household income groups (figure S1) and households with and without children (figure S2), compared to the counterfactual at 19 months post-implementation (Table S1).

### Purchasing of alcohol

Compared to the counterfactual estimated from pre-announcement trends, the volume of purchased alcohol reduced (P ≤0.05) overall and across all income groups (Figure S3) and in households with children (Figure S4). In households without children the volume of alcohol increased (P ≤0.05) compared to the predicted counterfactual (Table S2)

## Discussion

### Summary of principal findings

This is the first analysis to examine longer-term and differential impacts of the SDIL on changes in sugar from, and volume of, purchased soft drinks, and according to household income and composition. At 19 months post-implementation, sugar purchased from soft drinks fell overall (by 8g per household per week, or 3%) compared to the counterfactual of no intervention, but volume did not. Alongside, we found evidence that the SDIL reduced inequalities in sugar purchasing associated with soft drinks. Lower income households, and those with children, purchased the most sugar from, and volume of, soft drinks at baseline. They also had the largest reductions in sugar from (by 70g per household per week or 14% in the lowest income households and 56g or 14% in households with children), and volume of (by 675ml or 6%, and 849ml or 9% respectively), soft drinks purchased following the SDIL announcement and implementation. A 70g reduction in sugar per household per week is equivalent to just over two 250ml servings of a lower-levy tier drink per person per week, in an average UK household consisting of 2.4 people[42] than the decreases seen elsewhere. There was no evidence of substitution to confectionary or alcohol.

### Strengths and weaknesses

This study used nationally representative data on household purchases collected on a weekly basis over 295 weeks in a large sample. Availability of sociodemographic data enabled us to examine purchases by household income, a commonly used indicator of socioeconomic status[43], and presence of children at the household level. However, it was not possible to examine household composition in more granular detail due to limited data availability. The CITS analyses included a non-equivalent control category (toiletries) and at each time point accounted for important factors such as seasonal variations. We also explored the possibility of substitution with other potential sources of sugar (confectionary) and drinks (alcohol). With household purchasing data, it was not possible to record waste or the share of purchases among individuals within a household. The trajectories of the counterfactuals used in the CITS are modelled and based on the trends from March 2014 up until the SDIL announcement (March 2016). However, they may not have continued to take the same course. Attributing changes in the outcomes of interest to the SDIL requires consideration of other events, in particular the wider UK sugar reduction strategy. However, evidence so far suggests that the strategy has led to minimal changes in purchasing of sugar beyond the effects of the SDIL[44].

Rogers et al, previously found there was some evidence of a degree of error, but not bias, in some sugar concentrations reported by KWP compared with information provided on manufacturers’ websites[45].

### Comparison with other studies and interpretation of results

Sugar sweetened beverage taxes target whole populations, but potential health benefits may be greater for some population groups. Here, and in line with some previous studies [31,32,34] we observe that low income households were most responsive to the SDIL. Whilst others have hypothesised that this may be due to greater price sensitivity in lower income households, the SDIL had a complex impact on soft drinks prices[19]. Further, we find greater proportional drops in sugar than volume purchased, reinforcing the importance of reformulation alongside any individual level behaviour change. There were also marked differences in baseline purchasing with lower income households purchasing four times as much sugar from soft drinks compared to higher incomes ones. Thus, there may have been more room for lower income households to change their purchasing.

A novel element of our study (particularly timely given recent increases in childhood obesity in UK primary school children during the covid-19 pandemic [46]) was that households with children were more responsive to the SDIL than those without. Previous work has demonstrated that children are high consumers of SSBs[47] which, in turn, is associated with childhood obesity[10]. Furthermore the SDIL has been associated with a reduction in prevalence of obesity in girls aged 10/11 years in England[48] and a reduction in childhood hospital admissions for tooth extractions due to caries[49]. Our findings are compatible with previous studies showing that Mexican households with children reduced sugar from SSBs by 11% following introduction of an SSB tax, compared to only 2% in adult-only households[31]. As with lower-income households, greater responsiveness amongst households with children may be due to higher baseline purchasing and differential purchasing of drinks more likely to be reformulated. Furthermore, any signalling effect of the SDIL may have been more salient to households with children, particularly as it was part of the UK’s Childhood Obesity Plan[50].

Overall, we found that purchasing of sugar from soft drinks to reduced by 7.5g (2.6%) per household per week compared to the counterfactual at 19 months, whilst the volume purchased did not change. While many studies have reported reductions in purchases of taxed SSBs following implementation of taxes, the extent of the reductions differ considerably[51]. This likely reflects differences in the design of different taxes, differences in baseline consumption and the ease of citizens avoiding a tax by cross-border shopping. Many SSB taxes are intended to increase the price of SSBs relative to non-SSBs. In contrast, the SDIL was primarily intended to incentivise removal of sugar from drinks and this did occur[19]. The impact of the SDIL on prices was not straightforward, with price revisions across both levied and unlevied drinks [19]. As the SDIL was implementation nationwide, cross-border shopping is unlikely to be a significant concern. The overall effect size we found is similar to that found in relation to other tiered SSB taxes in Catalonia (2.2% reduction in sugar from SSBs) and in one study in Chile (3.4% in one study [52], although 21.6% was reported in another Chilean study[28]).

We find no evidence of a diminishing effect of SDIL on purchased sugar in soft drinks over time. Our findings of an overall reduction in weekly household sugar purchased form soft drinks of 7.5g (2.9g, 12.1g) compared to pre-announcement counterfactuals at 19-months post-implementation, are of a similar magnitude to analysis of similar data to 12 months follow up where a reduction in sugar of 8.0g (2.4g, 13.6g) was reported[18].This is consistent with findings of a non-diminishing influence of the tax on soft drinks in Mexico at 24 months post-tax implementation. Few studies have examined impacts beyond 12 months of a SSB tax being implemented[34,35,53].

Higher-income households, and those without children, on average showed slight increases in sugar purchased from soft drinks, compared to the counterfactual scenario. One explanation for this might be floor effects as these were the groups with the lowest levels of purchasing at baseline. It is also possible that these purchasers had preferences for drink products that did not undergo reformulation. It has also been observed that when facing government interventions some households respond counter-intuitively as a form of protest – termed psychological reactance. For instance, reactance was observed immediately following the referendum confirming an SSB tax in Berkeley[53]. In our study, small increases in purchasing of sugar from soft drinks are noticeable in high- and middle-income groups (in figures 2b and 2c) at the time of the SDIL announcement suggesting possible reactance.

Some studies have suggested that price changes in SSBs are linked to changes in purchasing of different alcoholic drinks[54]. We found little evidence that the SDIL increased purchasing of alcohol – indeed we found reductions in alcohol purchasing, compared to the counterfactual, in almost all demographic groups. Confectionery purchases remained stable with no evidence of substitution. This is in line with our previous study suggesting soft drinks were not substituted for by confectionery[18].

## Conclusions and implications for policy and research

Our findings suggest that the impact of the UK SDIL is likely to be greatest in the highest purchasing households (i.e. lower income households and those with children). Like other low-agency population interventions the SDIL, has the potential to decrease inequalities in dietary health. We also find persisting effects of the SDIL at 19 months post-implementation on purchasing of sugar from soft drinks, suggesting it may have longer term benefits for population dietary health. Small increases in sugar purchased from soft drinks were apparent in higher income households and households without children suggesting that a package of different interventions may be required to ensure all members of the population benefit from sugar reduction efforts.

## Funding

NR, MW, and JA were supported by the Medical Research Council (grant Nos MC_UU_00006/7). This project was funded by the NIHR Public Health Research programme (grant Nos 16/49/01 and 16/130/01). The views expressed are those of the authors and not necessarily those of the National Health Service, the NIHR, or the Department of Health and Social Care, UK. The funders had no role in study design, data collection and analysis, decision to publish, or preparation of the manuscript. NR had full access to all the data in the study and had final responsibility for the decision to submit for publication.

## Competing interests

All authors have completed the ICMJE uniform disclosure form at www.icmje.org/coi_disclosure.pdf (available on request from the corresponding author) and declare: MW is past director of the National Institute for Health Research Public Health Research Funding programme; no support from any organisation for the submitted work other than that described above; no financial relationships with any organisations that might have an interest in the submitted work in the previous three years; and no other relationships or activities that could appear to have influenced the submitted work.

## Ethical approval

Not required for secondary data analysis of anonymised data.

## Data sharing

Kantar Fast Moving Consumer Goods panel data are not publicly available but can be purchased from Kantar Worldpanel (http://www.kantarworldpanel.com). The authors are not legally permitted to share the data used for this study however interested parties can contact Kantar WorldPanel representative Sean Cannon to inquire about accessing this proprietary data.

The lead author affirms that the manuscript is an honest, accurate, and transparent account of the study being reported; that no important aspects of the study have been omitted; and that any discrepancies from the study as planned (and, if relevant, registered) have been explained.

Dissemination to participants and related patient and public communities: This work was presented at the 2022 annual scientific meeting of the Society of Social Medicine. We will issue a press release on this work and engage with media outlets as relevant. We will summarise our findings in a Twitter thread. A lay summary of this paper will be prepared in advance of publication and shared on the Medical Research Council Epidemiology Unit and Centre for Diet and Activity Research websites. We will share this summary with our networks of public health practitioners and policymakers through our social media accounts and regular e-newsletter. A lay summary of the findings of the wider project of which this is part will be made available on the National Institute for Health Research website.

Provenance and peer review: Not commissioned; externally peer reviewed.

## Supplementary Figures

**Figure S1:** Weight (g) of sugar from soft drink products purchased per household per week, by gross household income levels, from March 2014 to November 2019. Observed and modelled amounts of sugar in all soft drinks (drinks liable to the SDIL and non-liable drinks) by annual gross household income levels of a) <20,000 b) £20,000-£50,000 and c) £50,000 or more. Light blue points show observed data and light blue lines (with light blue shadows) shows modelled data (and 95% confidence intervals) of sugar from purchased soft drinks. The dark blue line indicates the counterfactual line based on preannouncement trends and had the announcement and implementation not happened. The first and second dashed lines indicate the announcement and implementation of SDIL, respectively. The red line (and shadow) indicates modelled toiletries (control group). The first and second dashed lines indicate the announcement and implementation of SDIL, respectively.

**Figure S2:** Weight (g) of sugar from soft drink products purchased per household per week, by whether households have children or not, from March 2014 to November 2019. Observed and modelled amounts of sugar in all soft drinks (drinks liable to the SDIL and non-liable drinks) by a) households with no children b) households with children (<16 years). Light blue points show observed data and light blue lines (with light blue shadows) shows modelled data (and 95% confidence intervals) of sugar from purchased soft drinks. The dark blue line indicates the counterfactual line based on preannouncement trends and had the announcement and implementation not happened. The first and second dashed lines indicate the announcement and implementation of SDIL, respectively. The red line (and shadow) indicates modelled toiletries (control group). The first and second dashed lines indicate the announcement and implementation of SDIL, respectively.

**Figure S3:** Weight (g) of sugar from confectionery purchased per household per week, by gross household income levels, from March 2014 to November 2019. Observed and modelled amounts of sugar in all soft drinks (drinks liable to the SDIL and non-liable drinks) by annual gross household income levels of a) <20,000 b) £20,000-£50,000 and c) £50,000 or more. Light blue points show observed data and light blue lines (with light blue shadows) shows modelled data (and 95% confidence intervals) of sugar from purchased soft drinks. The dark blue line indicates the counterfactual line based on preannouncement trends and had the announcement and implementation not happened. The first and second dashed lines indicate the announcement and implementation of SDIL, respectively. The scales on the Y axis varies between panels and modelled toiletries have been removed to maximise the resolution of the graphs

**Figure S4:** Weight (g) of sugar from confectionery purchased per household per week, by whether households have children or not, from March 2014 to November 2019. Observed and modelled amounts of sugar in all soft drinks (drinks liable to the SDIL and non-liable drinks) by a) households with no children b) households with children (<16 years). Light blue points show observed data and light blue lines (with light blue shadows) shows modelled data (and 95% confidence intervals) of sugar from purchased soft drinks. The dark blue line indicates the counterfactual line based on preannouncement trends and had the announcement and implementation not happened. The first and second dashed lines indicate the announcement and implementation of SDIL, respectively. Modelled toiletries have been removed (to maximise the resolution of the graphs.

**Figure S5:** Volume (mls) of alcohol products purchased per household per week, by gross household income levels, from March 2014 to November. Observed and modelled volumes of alcohol by annual gross household income levels of a) <20,000 b) £20,000-£50,000 and c) £50,000 or more. Light blue points show observed data and light blue lines (with light blue shadows) shows modelled data (and 95% confidence intervals) of volumes of alcohol. The dark blue line indicates the counterfactual line based on preannouncement trends and had the announcement and implementation not happened. The first and second dashed lines indicate the announcement and implementation of SDIL, respectively. The scales on the Y axis varies between panels and modelled toiletries have been removed (see figure S3 for inclusion of toiletries) to maximise the resolution of the graphs

**Figure S6:** Volume (mls) of alcohol purchased per household per week, by whether households have children or not, from March 2014 to November. Observed and modelled volumes of alcohol by a) households with no children b) households with children (<16 years). Light blue points show observed data and light blue lines (with light blue shadows) shows modelled data (and 95% confidence intervals) of volumes of alcohol. The dark blue line indicates the counterfactual line based on preannouncement trends and had the announcement and implementation not happened. The first and second dashed lines indicate the announcement and implementation of SDIL, respectively. The scales on the Y axis varies between panels and modelled toiletries have been removed (see figure S4 for inclusion of toiletries) to maximise the resolution of the graphs

**Table S1:** Absolute and relative changes in weight of sugar from confectionery, purchased per household per week compared to the counterfactual estimated from pre-announcement trends, at 19 months post-implementation of the UK soft drinks industry Levy.

| Sugar (g) | Pre-announcement to post implementation |  |
| --- | --- | --- |
|  | Absolute change (g) | Relative (%) |
| Total population | 6.40(-20.56, 33.36) | 3.14(-10.08, 16.36) |
| Income |  |  |
| <20,000 | -22.8 (-63.02, 17.41) | -7.07(-19.55, 5.40) |
| 20,000-50,000 | 10.05(-9.56, 29.65) | 6.73 (-6.40, 19.86) |
| >50,000 | 1.55(-17.94, 21.04) | 1.16(-13.41, 15.73) |
| Children |  |  |
| Yes | -15.50 (-46.39, 15.39) | -6.27(-18.77, 6.23) |
| No | 16.98(-9.64, 43.59) | 9.13(-5.18, 23.43) |

**Table S2:** Absolute and relative changes in volumes from alcohol purchased per household per week compared to the counterfactual estimated from pre-announcement trends, at 19 months post-implementation of the UK soft drinks industry Levy.

| Volume (mls) | Pre-announcement to post implementation |  |
| --- | --- | --- |
|  | Absolute change (ml) | Relative (%) |
| Total population<sup>1</sup> | -66.13 (-30.96, -101.31)* | -3.68 (-5.63, -1.72)* |
| Income |  |  |
| <20,000 | -177.55(-305.58, -49.52)* | -7.23(-12.44, -2.02)* |
| 20,000-50,000 | -84.83(-120.54, -49.11)* | -4.57(-6.50, -2.65)* |
| >50,000 | -93.7(-178.3, -9.12)* | -6.45(-12.28, -0.63)* |
| Children |  |  |
| Yes | -320.74(-411.22, -230.27)* | -18.66 (-23.93, -13.40)* |
| No | 81.55(45.92, 117.18)* | 4.61(2.59, 6.62)* |

<sup>1</sup>Whole population includes households with missing values for income (and which represents 8.2% of households) hence total population numbers may not reflect the range of values across income categories.

## Notes

### Competing Interest Statement

The authors have declared no competing interest.

### Summary of Updates

Figure Legends added Updated reference list included

