## Supplementary figures and images for "Changes in household purchasing of soft drinks following the UK Soft Drinks Industry Levy by household income and composition: controlled interrupted time series analysis, March 2014 to November 2019"

### Figure S1

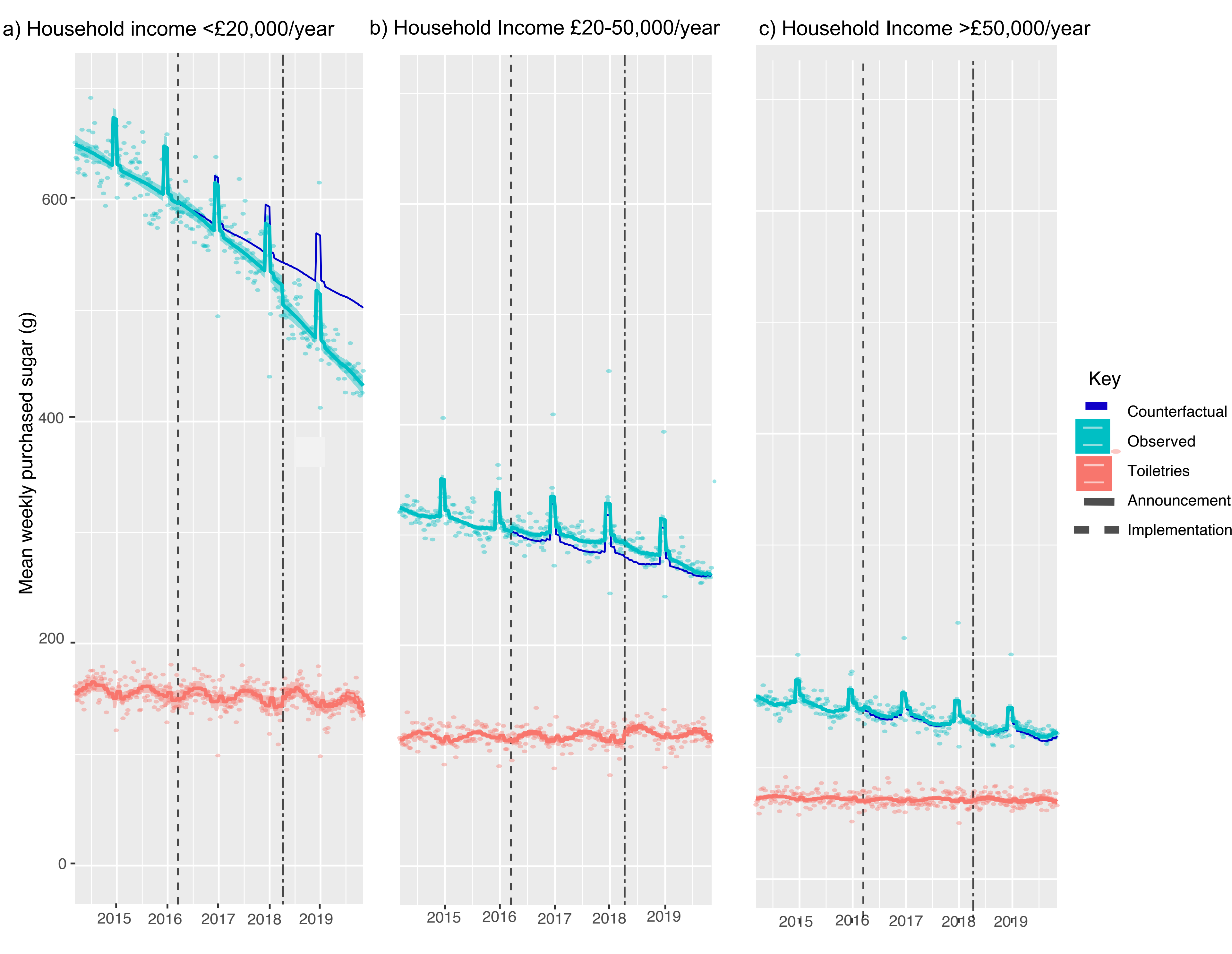

### Figure S2

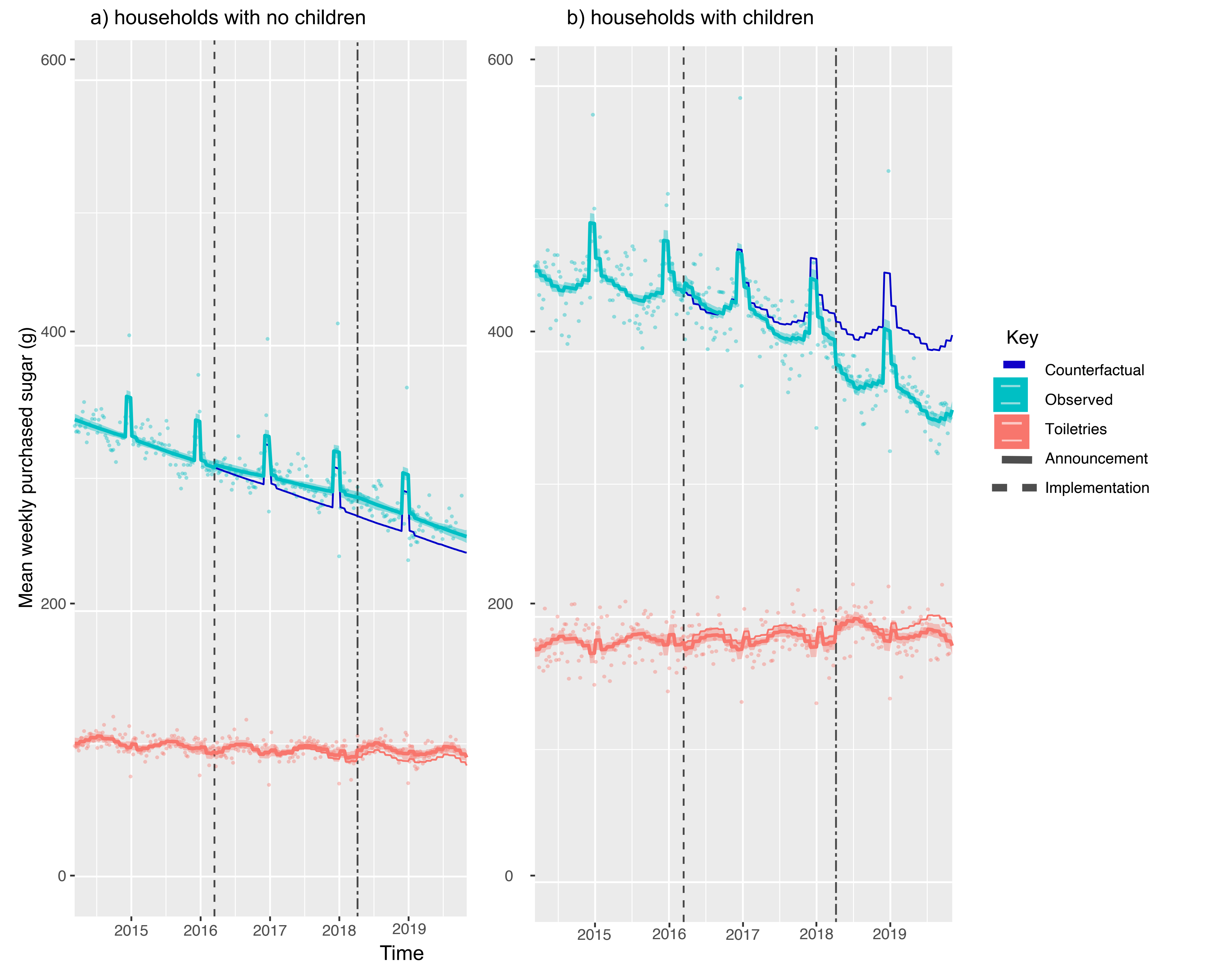

### Figure S3

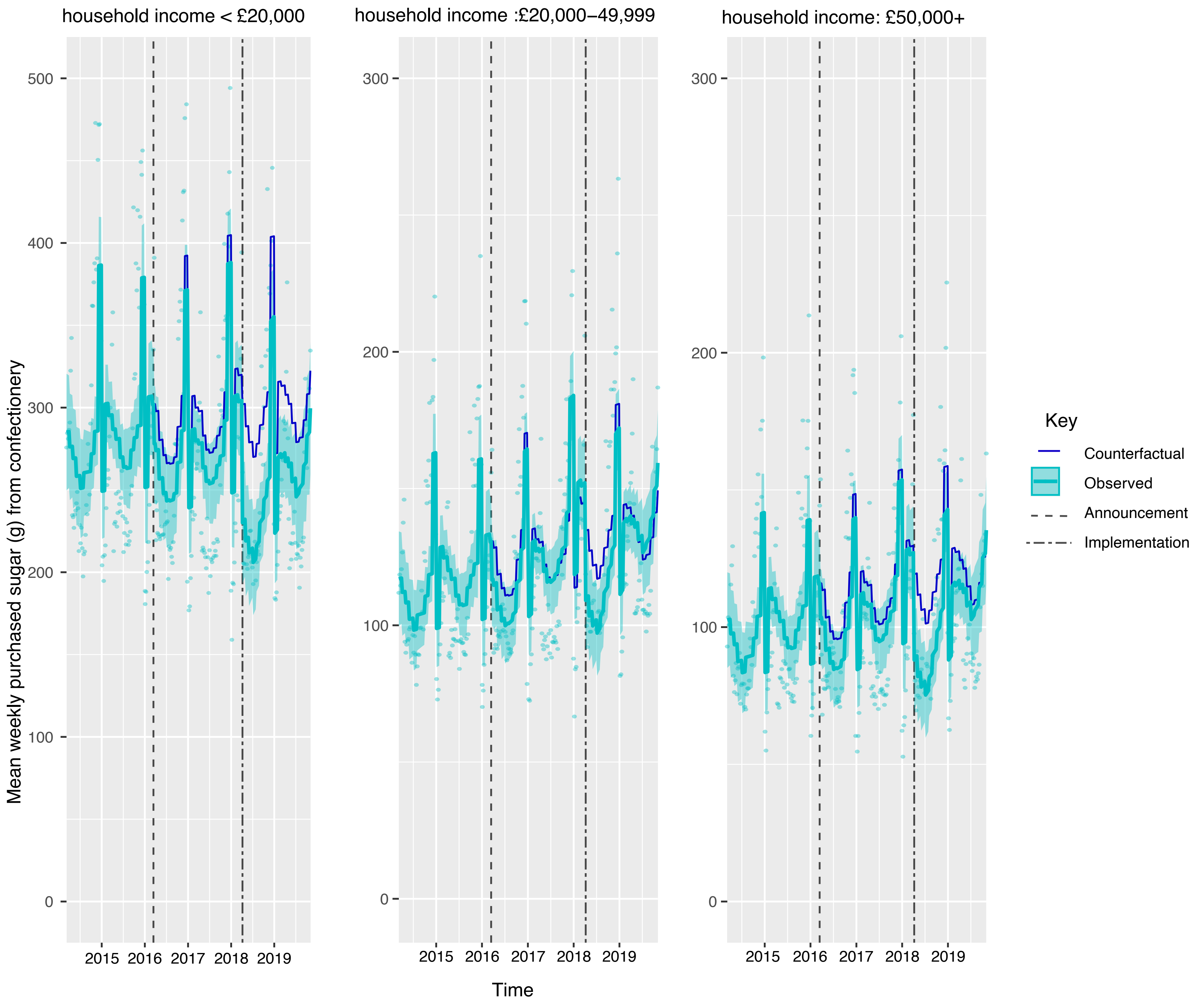

### Figure S4

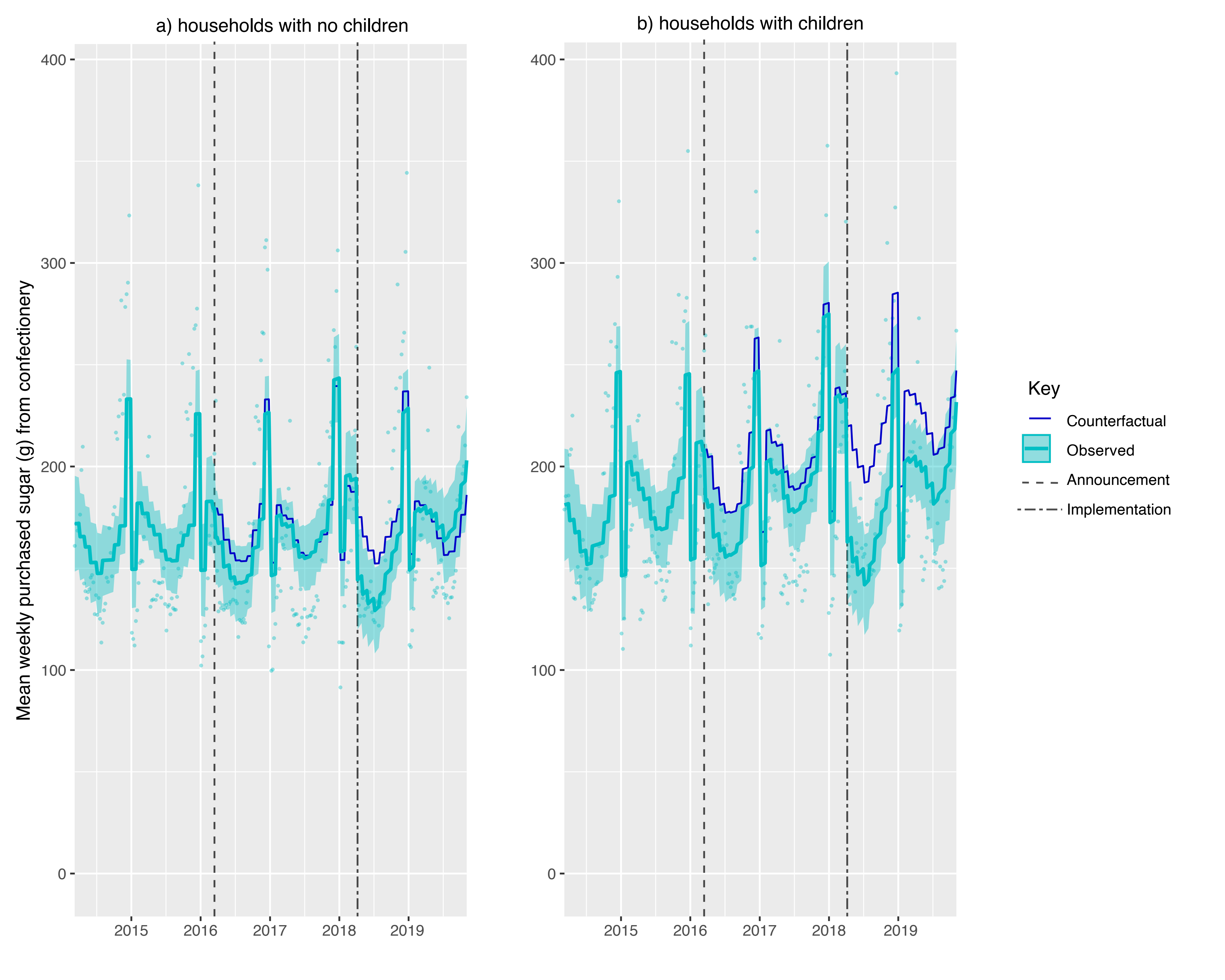

### Figure S5

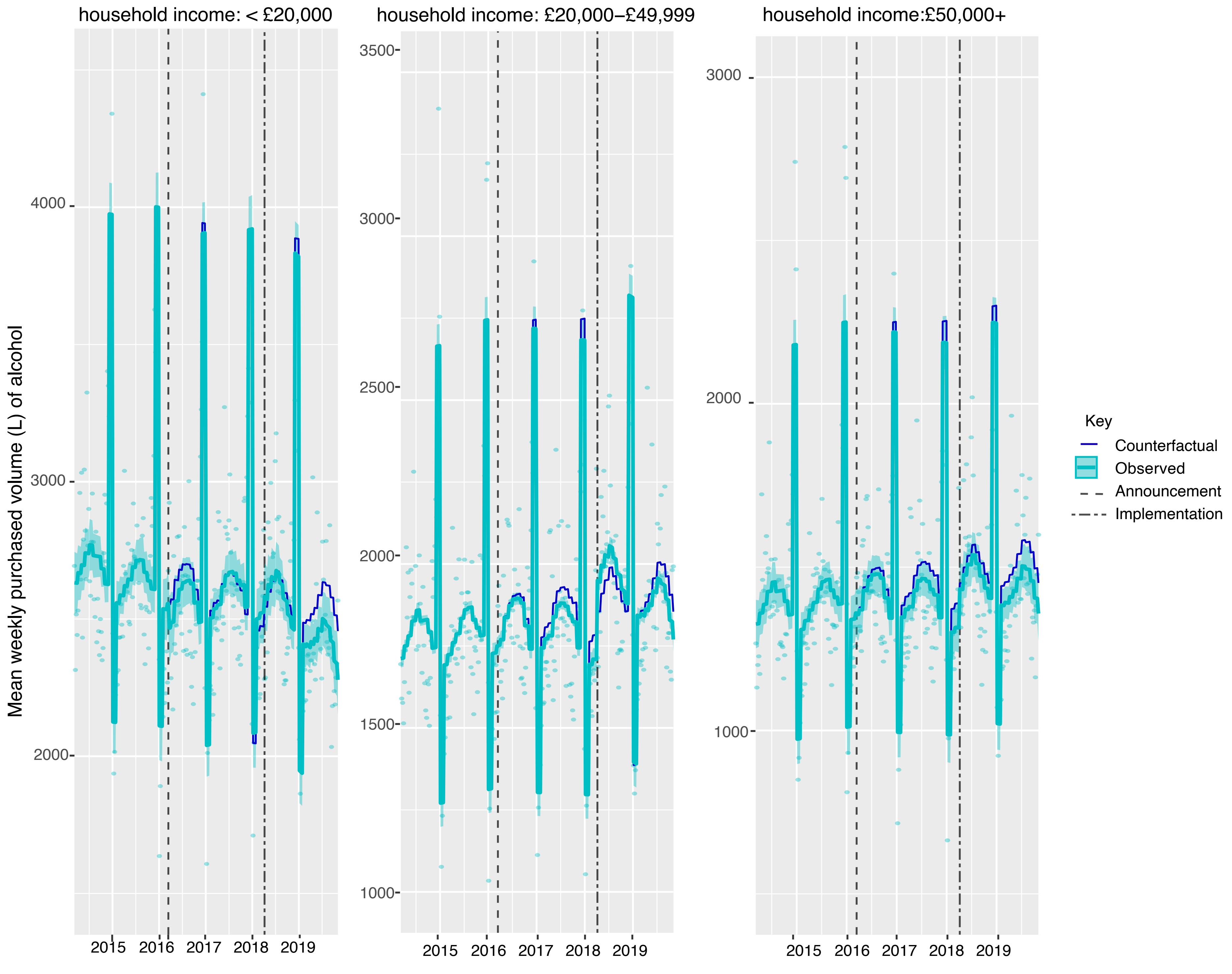

### Figure S6

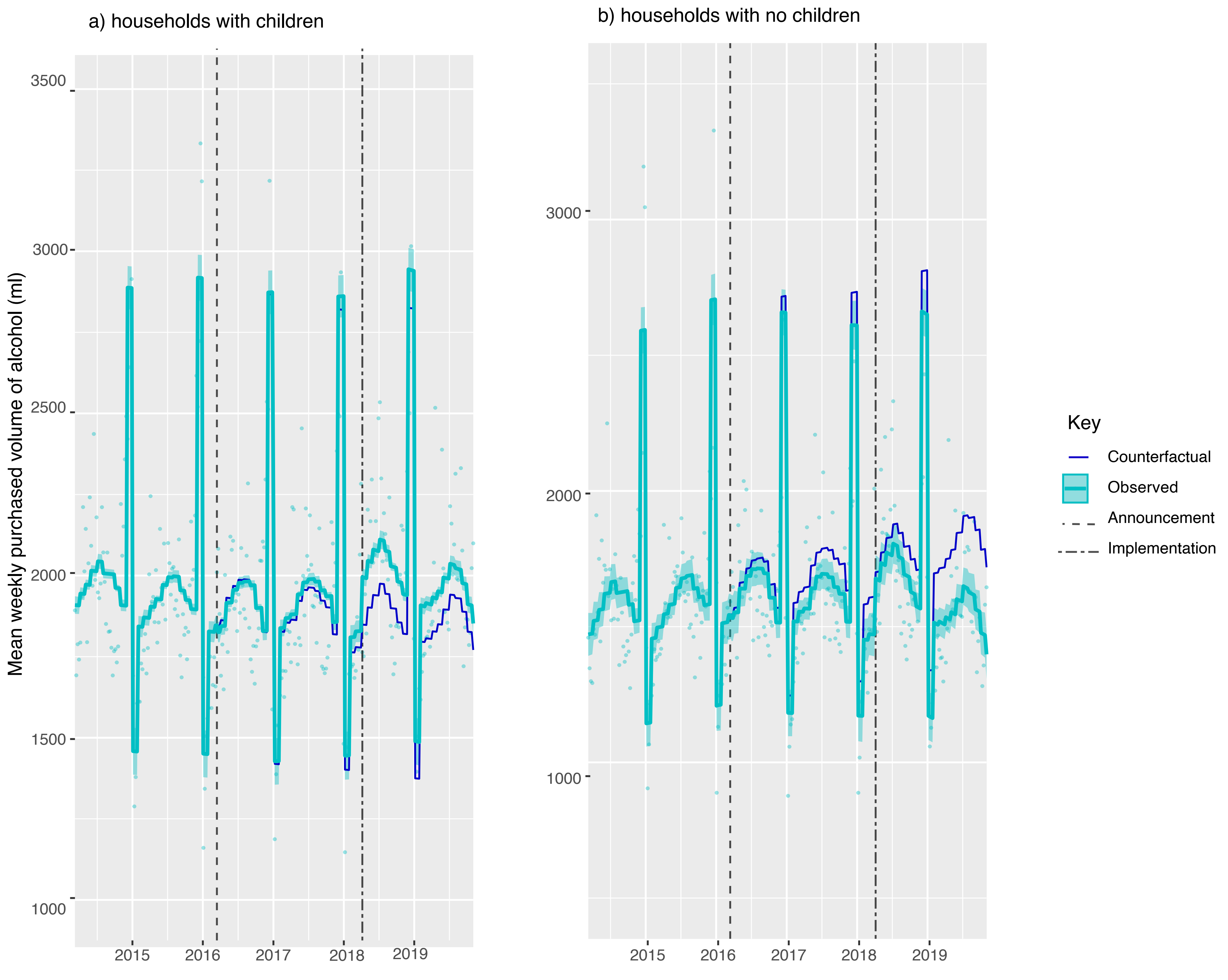
